# Predicting intention to vaccinate against COVID-19 in older Syrian refugees in Lebanon: findings from a multi-wave study

**DOI:** 10.1101/2022.07.23.22277948

**Authors:** Noura Salibi, Sawsan Abdulrahim, Maria El Haddad, Berthe Abi Zeid, Marwan F. Alawieh, Zeinab Ramadan, Hala Ghattas, Stephen J. McCall

## Abstract

**Introduction:** COVID-19 vaccine acceptance among refugees in the Arab region remains low. This study aimed to examine the prevalence, reasons and predictors of COVID-19 vaccine refusal among older Syrian refugees in Lebanon.

**Method:** A nested cross-sectional study among older Syrian refugees in Lebanon. The sampling frame was a complete listing of beneficiary households of a humanitarian organization with an adult aged 50 years or older. Telephone surveys were completed between September 2020 and May 2021. Logistic regression models were used to identify predictors of COVID-19 vaccine refusal. Models were internally validated using bootstrap methods and the models’ calibration and discrimination were presented.

**Results:** Of 3,173 Syrian refugees, 61% intended to receive the COVID-19 vaccine, 31% refused and 7% were undecided. Reasons for vaccine refusal were: preference to follow preventive measures (27%) and belief that the vaccine is not essential (21%). Despite high vaccine acceptance, only 6% of older Syrian refugees were registered on the national platform to receive the vaccine. Reasons for not registering included: being unsure about how to register (36%), and not wanting to receive the vaccine (33%). Predictors of COVID-19 vaccine refusal included: sex (female), older age, education, living outside informal tented settlements, perceiving COVID-19 as not severe and vaccines as not safe or effective, and using social media for information on COVID-19. After adjusting for optimization, the final model showed moderate discrimination (C-statistic: 0.65 (95% CI:(0.63-0.67)) and good calibration (C-Slope: 0.93 (95% CI:0.82-1.06)).

**Conclusion:** This study developed predictive model for vaccination intention with a moderate discriminative ability and good calibration. Prediction models in humanitarian settings can help to identify refugees at higher risk of not intending to receive the COVID-19 vaccine for public health targeting.

**What is already known on this topic:** Despite global efforts towards more inclusive national deployment vaccination plans, vaccine coverage and uptake among migrants and refugees remains low. Refugees and migrants, the majority of whom live in low and middle income countries, bear the double burden of vaccine inequity and face several challenges and barriers to vaccination including low vaccine supply, inability to access health services, fear of arrest and deportation, lack of accessible information as well as other language, and economic barriers. Research on COVID-19 vaccine intentions among refugees in the region has been limited. Understanding intentions and predictors to vaccinate among refugees, and addressing barriers to vaccine acceptance and registration, is crucial to ensure equitable vaccination and coverage, reduce the spread of COVID-19 and achieve herd immunity.

**What this study adds:** This study is one of the first to develop and internally validate a model of intention to refuse vaccination against COVID-19 in older Syrian refugees. Predictors of intention to refuse the vaccine include age, education, living outside informal tented settlements, sex, perceiving COVID-19 as not a serious infection and vaccines as not safe or effective, and using social media as a source of information on COVID-19. The primary reasons for vaccine refusal were: preference to follow preventive measures, concerns that the vaccine is too new, and belief that the vaccine is not essential. Registration on the national platform to receive the vaccine was low and the reasons for not registering included: being unsure about how to register, and not wanting to receive the vaccine.

**How this study might affect research, practice or policy:** This study highlights the need for targeted interventions to enhance vaccine acceptance and uptake among older Syrian refugees, and address barriers to vaccine registration. Predictors of COVID-19 vaccine refusal among older Syrian refugees will inform humanitarian programming and public health campaigns, and guide resource allocation and deployment planning. Findings inform future research to better understand the predictors of vaccine refusal.

## Introduction

The coronavirus disease (COVID-19) has exacerbated inequalities within and between countries and impacted the most vulnerable social groups including displaced populations who are estimated to exceed 84 million globally.^1 2^ Vaccination against COVID-19 is crucial to limit the spread of the virus and achieve herd immunity. Yet, by the end of August 2021, almost 57% of the population living in high-resource countries received the vaccine as compared to 2% in low-resource countries.^3^

Access to life-saving vaccines vary among different population groups and a complex constellation of factors particularly impact access for migrants and refugees. ^4–7^ A review conducted by the World Health Organization (WHO) on 104 national deployment vaccination plans (NDVPs) showed that migrants were not explicitly included in 72% of NDVPs. Despite global efforts towards more inclusive NDVPs, actual implementation and vaccine coverage for migrants remain far from universal.^8^ Besides, in many countries, most vaccination campaigns only include migrants with regular documentation while those without documentation are left behind.^9^ Moreover, refugees, 86% of whom live in low- and middle-income countries, bear the double burden of vaccine inequity and face several challenges and barriers to vaccination including low vaccine supply, inability to access health services, fear of arrest and deportation, lack of accessible information, as well as other language and economic barriers.^4–7^

The Arab region hosts over 40 million migrants and refugees.^10^ Lebanon, a country enmeshed in ongoing political, social, and economic crises, hosts the highest number of refugees per capita.^11^ The country’s response to the COVID-19 pandemic was impacted by this context and the Beirut port explosion on August 4, 2020 which placed added strains on an already overwhelmed, and highly privatized health care system. In 2021, there was an estimated 855,000 Syrian refugees in Lebanon;^12^ of those, only 16% percent aged 15 years and above had legal residency and 90% were living in extreme poverty^13^. Thus, Syrian refugees are particularly vulnerable to the direct and indirect effects of COVID-19.

Research on COVID-19 vaccine intentions among refugees and migrants in the region has been limited. Existing evidence from a study conducted among Qatari nationals and migrants between October and November, 2020 showed that around 16.7% of migrants were not willing to take the vaccine. Concerns about COVID-19 vaccine’s safety and long-term side effects were the main reasons for vaccine refusal.^14^

In Lebanon, despite the rapid spread of the COVID-19 virus since the first wave in February 2020, no outbreak was recorded among Syrian refugees in the country; yet, this might not be accurate as testing has remained low among Syrian refugees and national data on the prevalence of COVID-19 among this population is unavailable. ^15^ The Lebanon NDVP was launched in January 2021 and pledged to provide the vaccine for free to everyone on Lebanese soil irrespective of nationality or residency status.^16^ In collaboration with the Lebanese Ministry of Public Health (MOPH), the COVAX platform was developed to digitize registering for and receiving the COVID-19 vaccine.^16^ However, up to January 2022, vaccine coverage and uptake among refugees remained low with only 14% of Syrian refugees having registered for the COVID-19 vaccine on the national platform as compared to 79% of Lebanese. Similarly, 8% of the vaccine doses administered were for Syrians as compared to 85% for Lebanese.^17^

Understanding the predictors of COVID-19 vaccine acceptance and refusal among refugees is crucial to addressing barriers and ensuring equitable vaccination coverage, reducing the spread of COVID-19, and achieving herd immunity. As intention to adopt a behavior correlates with the behavior itself^18^, we examined in this study older Syrian refugees’ intentions to accept or refuse the COVID-19 vaccine and assessed the predictors of vaccine refusal among this vulnerable group in Lebanon.

## Materials and Methods

### Study design and setting

This was a nested cross-sectional study within a larger longitudinal study that aimed to track older Syrian refugees’ vulnerability to COVID-19 over time. The sampling frame included a complete listing of beneficiary households of a humanitarian organization that had at least one adult known to be 50 years or older, residing in tented settlements or localities in Lebanon, and who received humanitarian assistance during 2017-2021. Respondents participated in a telephone interview at months 1 starting in September 2020 (wave1), months 2 (wave 2), months 5 (wave 3), months 6 (wave 4) and months 17 (wave 5). This study will examine vaccine intention at a single point in time at wave 3 and vaccine registration at wave 4. The preliminary findings of this study have been previously published on a subset of the study population.^19^

### Study Population

The study population was Syrian refugees aged 50 years or older who participated in a multi-wave longitudinal study. 17,384 Syrian households in Lebanon that received assistance from a humanitarian organization during 2017 to 2020 were contacted, screened for eligibility and assessed for capacity to consent. Intention to receive the COVID-19 vaccine was collected in wave 3 (January 2021 – April 2021) and vaccination registration on the Lebanese platform was collected in wave 4 (February 2021 – May 2021).

### Study instrument and data collection

The study instrument for the longitudinal study was developed using existing validated scales and new questions through co-creation including input from academics, the humanitarian partner, government representatives, and refugee focal points. The instrument included 17 modules including basic socio-demographics, health, COVID-19, shelter, household water insecurity, safety and security, social support, violence and trauma, decision making, communication, assets, expenditure, assistance, income, debt, food security, and regularization. Building on the existing literature, culturally relevant questions were included as well as validated scales (Household Water Insecurity Experiences 4-item (HWISE-4) Scale,^20^ and Food Insecurity Experience Scale (FIES)^21^).

### Patient and public participation

Prior to data collection, community consultations were conducted in order to better understand the acceptability of the study and validity of the study instrument. Based on the community feedback and suggestions, the survey tool was adjusted to capture contextually-relevant perceptions and experiences that were not previously included. These consultations were used to inform the research project, and improve protection outcomes in humanitarian programming.

The survey tool was developed in English and translated to Arabic. The Arabic version of the instrument was pilot tested among a sample of Syrian refugees for face validity. Data collectors were trained to undertake the telephone interview and data entry checks were undertaken to ensure complete and reliable data.

### Outcome Measures

The main study outcome was intention to refuse the COVID-19 vaccine. As the Lebanon COVID-19 vaccine campaign’s launch in February 2021 coincided with the study’s wave 3, a question on intention to receive the COVID-19 vaccine was added to the study instrument. This question, the present study’s primary outcome, was assessed as follows: “If a safe and effective vaccine for COVID-19 became available, for free, would you take it?”. When the COVID-19 vaccine became available in Lebanon, this question was rephrased during wave 3 to become: “Now that a safe and effective COVID-19 vaccine arrived to Lebanon and is offered for free, would you take it”. Participants who answered No on any of these two questions were categorized as intending to refuse the vaccine.

Data on actual registration for the COVID-19 vaccine were collected one month later in wave 4 using the following question: “Have you (or someone on your behalf) registered for the COVID-19 vaccine?”.

### Candidate predictors

A list of 15 candidate predictors was included based on the literature on vaccine acceptance/refusal. These were: 1) socio-demographic variables: age; sex (female/male); residence outside or inside informal tented settlements (ITS); education (preparatory and higher/elementary/never attended); and employment status (employed/unemployed); 2) health related questions on the presence of chronic conditions (none/at least one), hypertension (yes/no), and diabetes (yes/no); 3) general perceptions towards vaccine safety and effectiveness; 4) COVID-19 related questions on adherence measures such as attended social events (yes/no), mainly stayed at home except for essential purchasing (yes/no), received visitors at home (yes/no), and worn a mask (yes/no), and on COVID-19 perceived severity (true/false) and susceptibility (yes/no); 5) Receipt of cash assistance (yes/no); and 6) Use of social media as an information source on COVID-19 (yes/no).

### Ethical approval

This study was granted approval by the Social and Behavioral Sciences Institutional Review Board (SBS-IRB) at the American University of Beirut. Oral consent was obtained from all participants in the study.

### Statistical Analysis

Simple frequencies were conducted for descriptive statistics. Unadjusted logistic regression models were run to examine the association between COVID-19 vaccine refusal and socio-demographic factors, COVID-19 adherence practices, and general vaccine as well as specific COVID-19 related perceptions.

A multicollinearity test was performed with a variance inflation factor greater than 5 to indicate collinearity. Vaccines’ safety and effectiveness were collinear and were modeled separately. Missing values did not exceed 5% for all included predictors. All variables were categorical except for age, which had a linear relationship with COVID-19 vaccine refusal. The model was built using backward multivariable logistic regression (P-Value: <0.157) with all candidate predictors of vaccine refusal to examine the prediction model.

The model’s performance was assessed in terms of discrimination using the C-statistic ranging between 0.5 and 1.0. The model calibration was assessed using calibration plot and C-slope; a slope of 1 and intercept of 0 shows perfect calibration or agreement between observed outcomes and predictive probabilities while a slope less than 1 is indicative of model overfitting. Calibration-in-the-large was presented to understand the agreement between predictive risk and observed events.

The final model was validated using bootstrap methods; 500 bootstrap samples with replacement were drawn to provide an estimate of the model’s optimism. An adjusted C-statistic and calibration plots were generated. To adjust for overfitting, bootstrap shrinkage was applied and an optimism adjusted odds ratios and beta coefficients were generated. A sensitivity analysis was conducted where perceptions of the effectiveness of vaccine was included in the model development process instead of perceptions of vaccine safety. All analyses were conducted using STATA/SE 17.

## Results

A total of 4,010 participants in these households were eligible and invited to participate, of whom 3,838 consented and participated in wave 1. Of this original sample, 3,173 participated in wave 3 and 2,990 participated in wave 4 (figure 1). Among those who participated in wave 3, n=3,173 had available data on vaccine intention. Additional data on vaccine registration was available for n=2,990 among those who participated in wave 4 sample. The median age of participants was 56 years (IQR: 53-63), of these 47.2% were females and the majority (62.2%) lived outside ITS (Table 1).

**Table 1.** **Key characteristics, beliefs, knowledge and medical history and their associations with intention to vaccinate against COVID-19**

|  | Total<br>(n=3,173)<br>n (%) | Intention to vaccinate<br>(n=1,941 (61.3%))<br>n (%) | Intention to refuse<br>the vaccine<br>(n=993 (31.3%))<br>n (%) | Undecided<br>(n=233 (7.4%))<br>n (%) | Intention to<br>refuse the<br>vaccine<sup>a</sup><br>OR (95% CI) |
| --- | --- | --- | --- | --- | --- |
| **Socio- Demographics** |  |  |  |  |  |
| *Age (years) (median (IQR))* | 3,173 (56 (53-63)) | 1,941 (56 (52-62)) | 993 (57 (53-64)) | 233 (58 (54-64)) | 1.01 (1.00-1.02) |
| **Sex** |  |  |  |  |  |
| Male | 1,674 (52.8) | 1,084 (64.9) | 479 (28.7) | 107 (6.4) | 1 |
| Female | 1,499 (47.2) | 857 (57.3) | 514 (34.3) | 126 (8.4) | 1.36 (1.16-1.58) |
| **Residence** |  |  |  |  |  |
| Inside ITS | 1,198 (37.8) | 766 (64.1) | 339 (28.4) | 89 (7.5) | 1 |
| Outside ITS | 1,975 (62.2) | 1,175 (59.6) | 654 (33.1) | 144 (7.3) | 1.26 (1.07-1.47) |
| **Education** |  |  |  |  |  |
| Never attended school | 1,884 (49.2) | 938 (61.3) | 467 (30.5) | 125 (8.2) | 1 |
| Elementary | 970 (25.4) | 511 (62.6) | 257 (31.5) | 48 (5.9) | 1.01 (0.84-1.22) |
| Preparatory and higher | 973 (25.4) | 487 (59.9) | 266 (32.7) | 60 (7.4) | 1.10 (0.91-1.32) |
| **Health** |  |  |  |  |  |
| **Chronic conditions** |  |  |  |  |  |
| None | 884 (28.9) | 568 (64.3) | 257 (29.1) | 58 (6.6) | 1 |
| At least one | 2,177 (71.1) | 1,304 (60.1) | 702 (32.3) | 166 (7.6) | 1.19 (1.00-1.41) |
| **Hypertension** |  |  |  |  |  |
| No | 1,759 (57.9) | 1,090 (62.0) | 541 (30.8) | 126 (7.2) | 1 |
| Yes | 1,281 (42.1) | 772 (60.5) | 408 (31.9) | 97 (7.6) | 1.06 (0.91-1.25) |
| **Diabetes** |  |  |  |  |  |
| No | 2,342 (76.8) | 1,445 (61.8) | 728 (31.1) | 166 (7.1) | 1 |
| Yes | 709 (23.2) | 421 (59.6) | 228 (32.2) | 58 (8.2) | 1.07 (0.89-1.29) |
| **Vaccination Perceptions** |  |  |  |  |  |
| **I think vaccines are safe** |  |  |  |  |  |
| Agree | 2,273 (71.6) | 1,561 (68.8) | 596 (26.3) | 113 (5.0) | 1 |
| Neither agree nor disagree or Don't know | 548 (17.3) | 280 (51.2) | 186 (34.0) | 81 (14.8) | 1.74 (1.41-2.14) |
| Disagree | 352 (11.1) | 100 (28.6) | 211 (60.3) | 39 (11.1) | 5.53 (4.28-7.14) |
| **I think vaccines are effective** |  |  |  |  |  |
| Agree | 2,295 (72.3) | 1,580 (68.9) | 601 (26.2) | 111 (4.8) | 1 |
| Neither agree nor disagree or Don't know | 574 (18.1) | 285 (49.7) | 201 (35.1) | 87 (15.2) | 1.85 (1.51-2.27) |
| Disagree | 304 (9.6) | 76 (25.2) | 191 (63.2) | 35 (11.6) | 6.61 (4.98-8.76) |
| **COVID-19 Adherence Measure** |  |  |  |  |  |
| **Attended social events** |  |  |  |  |  |
| No | 2,887 (94.3) | 1,766 (61.3) | 905 (31.4) | 210 (7.3) | 1 |
| Yes | 174 (5.7) | 104 (59.8) | 56 (32.2) | 14 (8.0) | 1.05 (0.75-1.47) |
| **Mainly stayed at home except for essential purchasing** |  |  |  |  |  |
| No | 702 (22.9) | 434 (61.9) | 213 (30.4) | 54 (7.7) | 1 |
| Yes | 2,362 (77.1) | 1,439 (61.1) | 748 (31.7) | 170 (7.2) | 1.06 (0.88-1.28) |
| **Received visitors at home** |  |  |  |  |  |
| No | 2,051 (67.0) | 1,249 (61.0) | 645 (31.5) | 153 (7.5) | 1 |
| Yes | 1,012 (33.0) | 623 (61.7) | 316 (31.3) | 71 (7.0) | 0.98 (0.83-1.16) |
| **Worn a mask** |  |  |  |  |  |
| No | 294 (9.6) | 175 (59.5) | 102 (34.7) | 17 (5.8) | 1 |
| Yes | 2,769 (90.4) | 1,698 (61.4) | 858 (31.1) | 207 (7.5) | 0.87 (0.67-1.12) |
| **COVID-19 Perceived severity** |  |  |  |  |  |
| **COVID-19 is a serious infection that is spreading across the world** |  |  |  |  |  |
| True | 3,015 (95.0) | 1,863 (61.9) | 925 (30.7) | 221 (7.3) | 1 |
| False | 158 (5.0) | 78 (49.4) | 68 (43.0) | 12 (7.6) | 1.75 (1.26-2.45) |
| **COVID-19 Perceived susceptibility** |  |  |  |  |  |
| No | 693 (21.8) | 401 (57.9) | 243 (35.1) | 49 (7.1) | 1 |
| Yes | 2,221 (70.0) | 1,387 (62.5) | 669 (30.2) | 161 (7.3) | 0.79 (0.66-0.96) |
| Don't know | 259 (8.2) | 153 (59.5) | 81 (31.5) | 23 (9.0) | 0.87 (0.64-1.19) |
| **Cash Assistance** |  |  |  |  |  |
| Yes | 2,876 (90.8) | 1,764 (61.5) | 895 (31.2) | 211 (7.3) | 1 |
| No | 293 (9.2) | 175 (59.7) | 96 (32.8) | 22 (7.5) | 1.08 (0.83-1.40) |
| **COVID-19 source of information** |  |  |  |  |  |
| **Use of social media** |  |  |  |  |  |
| No | 1,538 (48.5) | 994 (64.8) | 449 (29.2) | 92 (6.0) | 1 |
| Yes | 1,635 (51.5) | 947 (58.0) | 544 (33.3) | 141 (8.7) | 1.27 (1.09-1.48) |

<sup>a</sup>The unadjusted ORs are for refusal of COVID-19 vaccine versus acceptance; the "undecided" were excluded from logistic regression analyses.

**Figure 1.**
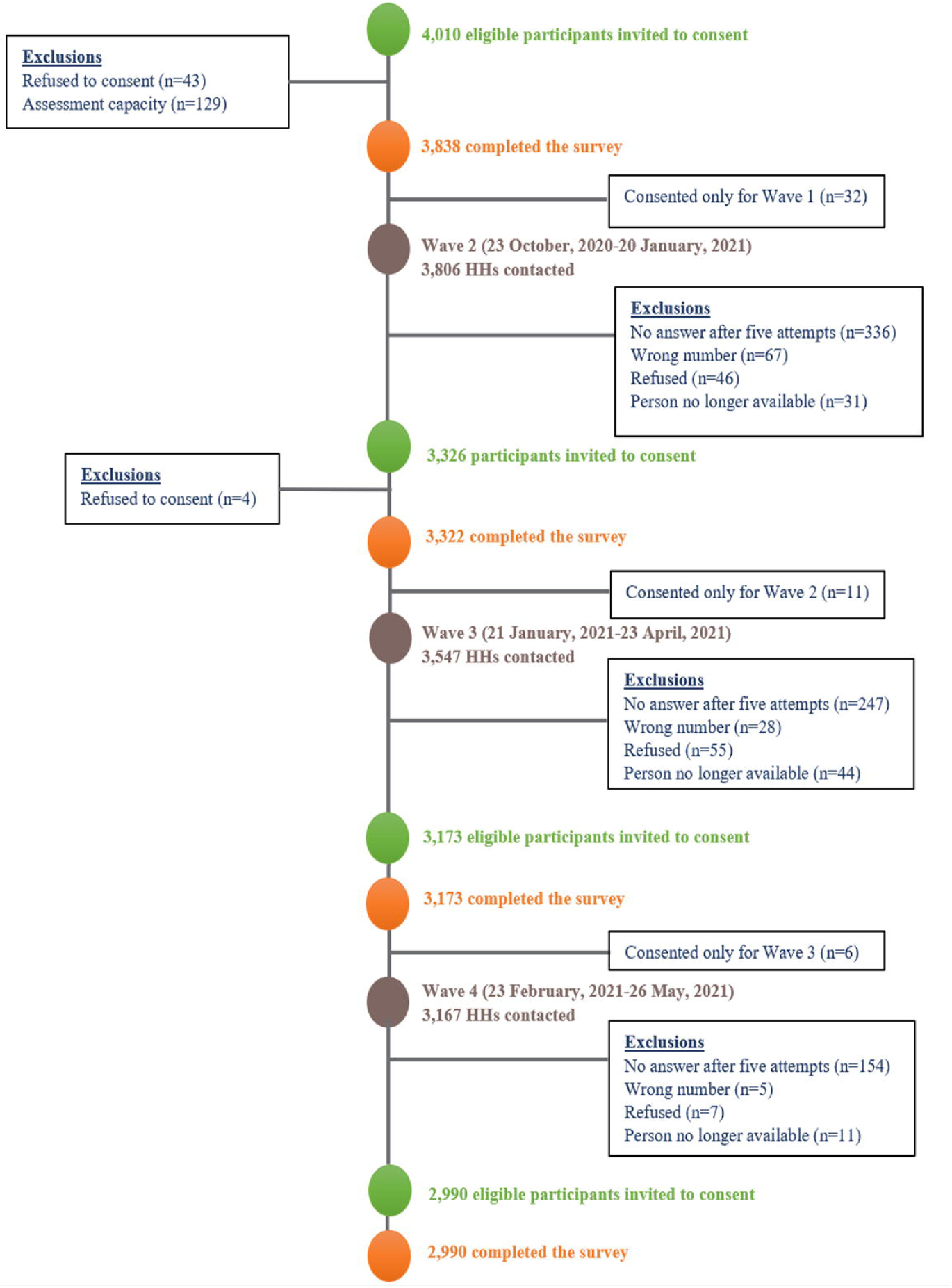
Flowchart of sample selection across waves.

### Preference for COVID-19 vaccine

Of 3,173 older Syrian refugees, 61.3% reported that they intend to receive the vaccine, 31.3% reported refusal, and 7.4% were undecided (Table 1). Reasons for vaccine refusal include preference to continue following COVID-19 precaution measures rather than take the vaccine (27.4%), concerns that the vaccine is too new and preference to wait until more is known (23.5%), belief that the vaccine is not essential (20.7%), and concerns regarding the vaccine’s side effects (10.1%) (Figure 2). Other reasons that were reported less frequently include: not believing that COVID-19 exists or perceiving that it does not require a vaccine, fear of needles, lacking trust in the system, believing that enough immunity was gained through previous COVID-19 infection, or perceiving that the vaccine should not be taken in the presence of allergies or other comorbidities (Figure 2).

**Figure 2.**
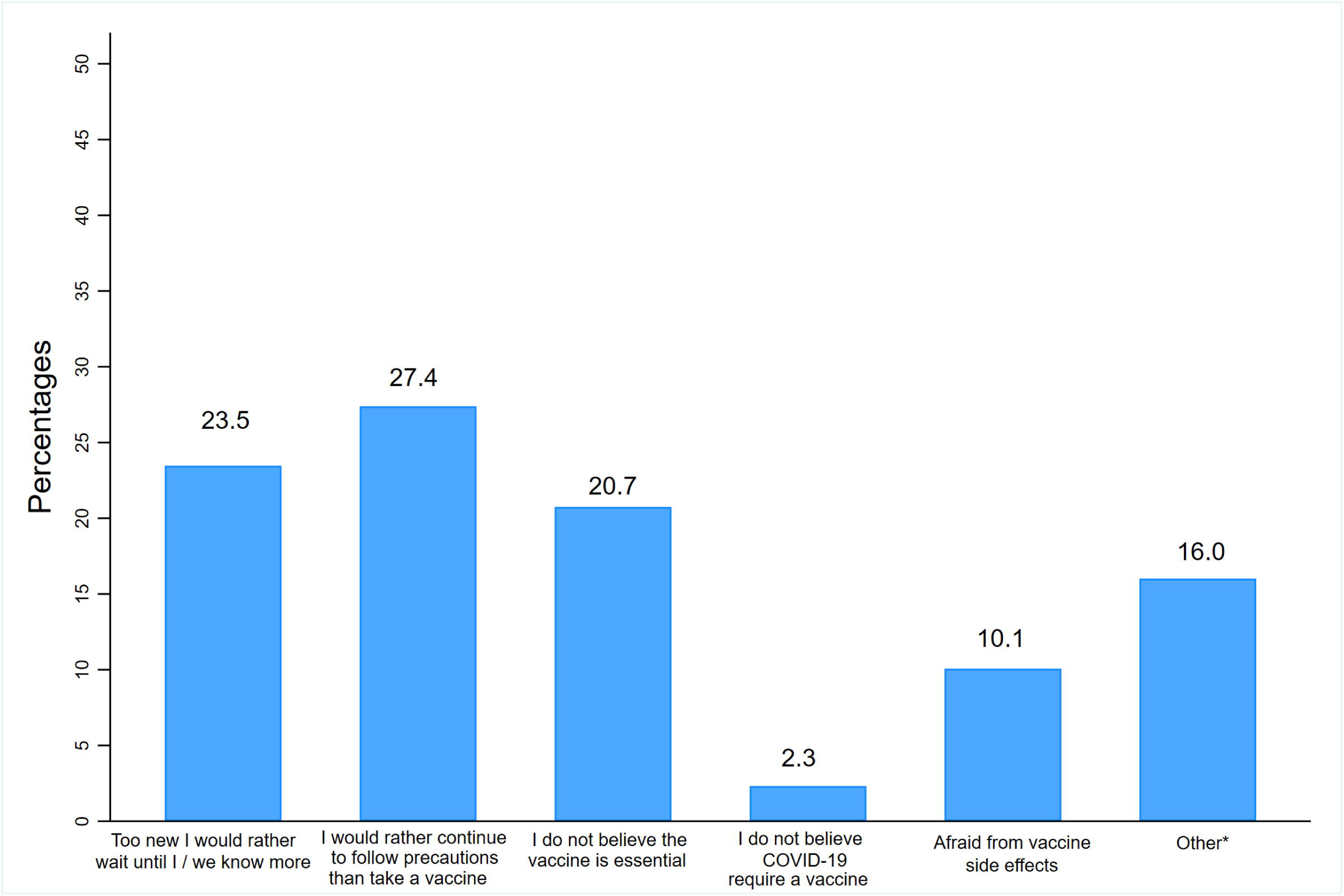
Reasons for intentions to refuse the COVID-19 vaccine (n=993).

**Figure 3a.**
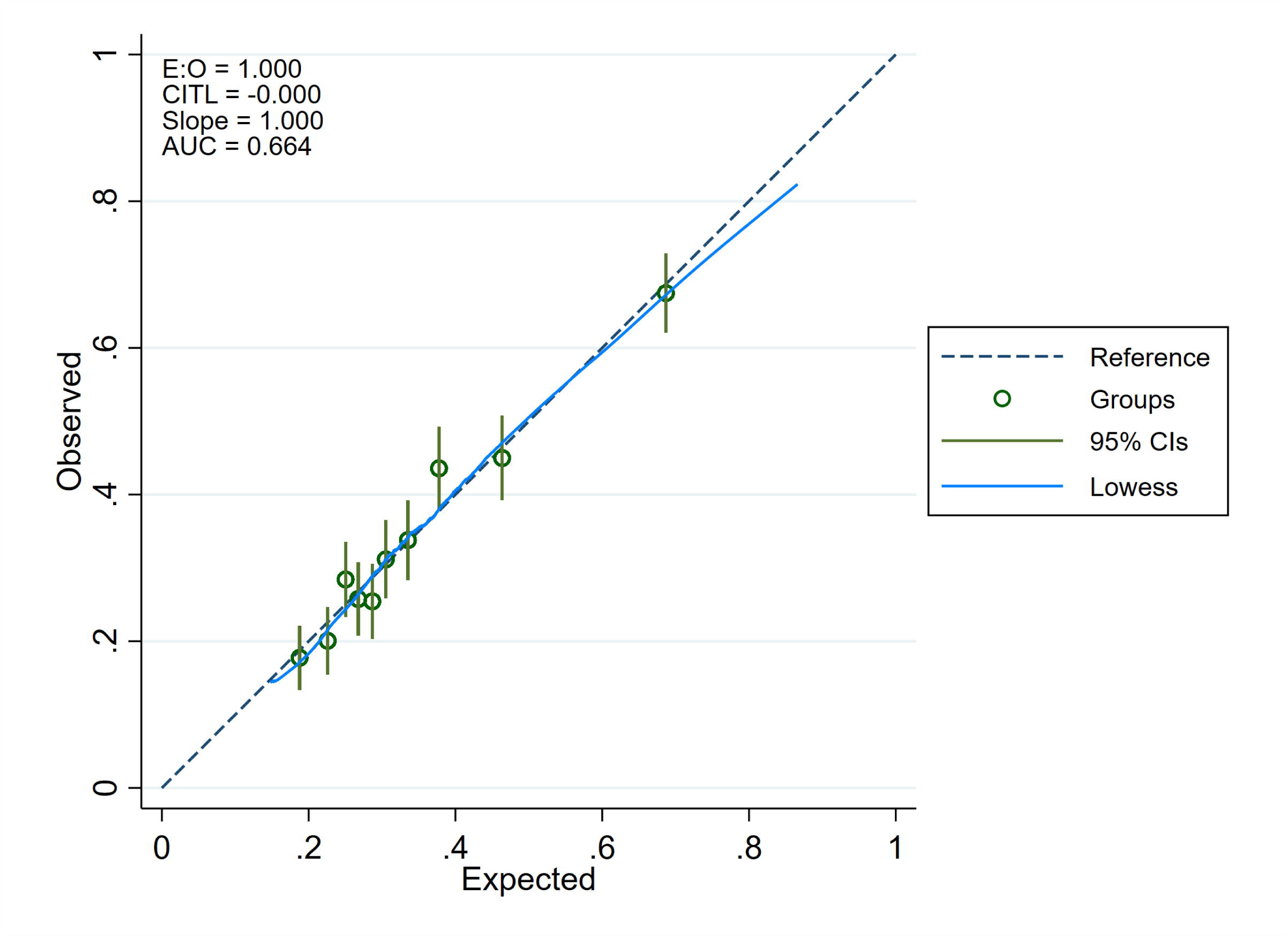
Model performance for the apparent model.

**Figure 3b.**
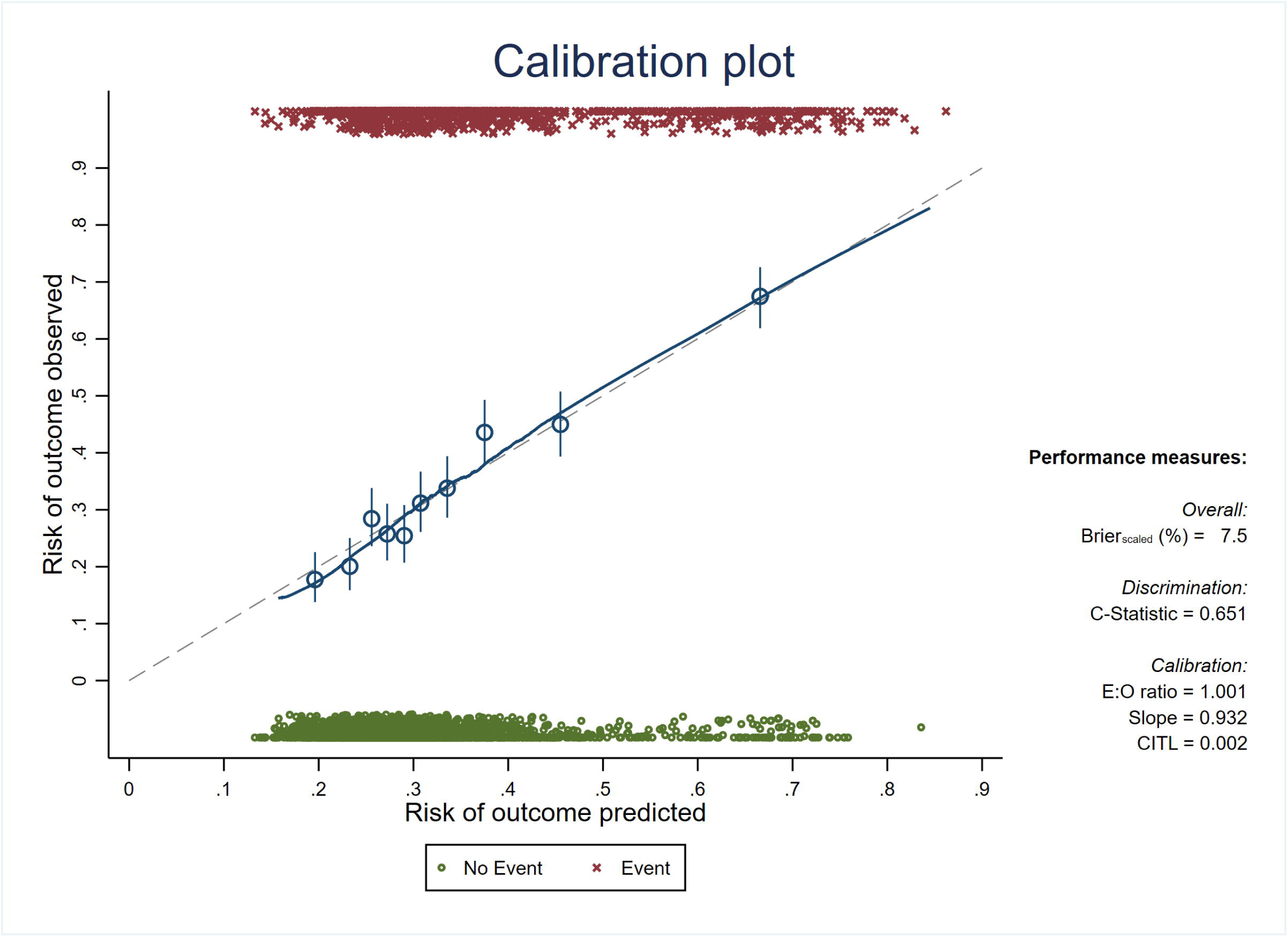
Model performance after adjusting for optimism.

### Bivariable analysis of study variables by COVID-19 vaccine refusal

In unadjusted analysis, females were more likely to report vaccine refusal as compared to males [OR:1.36; (95% CI:1.16-1.58)], and refugees living outside ITS compared those living inside ITS [OR:1.26; (95% CI:1.07-1.47)] (Table 1). In addition, those who perceived that vaccines are not safe [OR:5.53; (95% CI:4.28-7.14)] or not effective [OR:6.61; (95% CI:4.98-8.76)] were six times more likely to report vaccine refusal as compared to those who perceived them to be safe and effective (Table 1).

Although the majority (95%) of participants agreed with the statement that *‘COVID-19 is a serious infection that is spreading around the world’*, those who disagreed with it were more likely to report vaccine refusal as compared to those who agreed [OR: 1.75; (95% CI: 1.26-2.45)]. Moreover, although 22% of refugees did not perceive themselves to be susceptible to COVID-19 infection, those who perceived themselves susceptible to COVID-19 [OR:0.79; (95% CI: 0.66-0.96)] were less likely to refuse the COVID-19 vaccine. In addition, participants who used social media as a source of information on COVID-19 were more likely to report COVID-19 vaccine refusal as compared to those who did not use social media as a source of information [OR: 1.27; (95% CI: 1.09-1.48)].

### Predictors of COVID-19 vaccine refusal intention

Predictors of COVID-19 vaccine refusal were: age, sex, residence, education, perceptions around COVID-19 as a severe illness, perception about vaccine safety, and use of social media as a source of information (Table 2). The apparent model had a C-statistic of 0.66 (95% CI: 0.64-0.69) indicating moderate discrimination ability of the model. After adjusting for optimism, the C-statistic was 0.65 (95% CI: 0.63-0.67) and the calibration slope showed minor evidence of model overfitting (C-slope: 0.93 (95% CI:0.82-1.07). The calibration-in-the-large was 0.002 (95% CI: −0.08-0.09) (Table 2).

**Table 2.** Multivariable model for predicting intentions to refuse the COVID-19 vaccine.

|  | Apparent Model |  |  |  |  | Model adjusted by bootstrap shrinkage |  |  |  |  |
| --- | --- | --- | --- | --- | --- | --- | --- | --- | --- | --- |
|  | Parameter estimate | 95%CI | Odds Ratio | 95% CI | P-value | Parameter estimate | 95%CI | Odds Ratio | 95% CI | P-value |
| **Age** | 0.02 | 0.01-0.03 | 1.02 | 1.01-1.03 | 0.004 | 0.01 | 0.00-0.02 | 1.01 | 1.00-1.02 | 0.004 |
| **Sex** |  |  |  |  |  |  |  |  |  |  |
| Male | Ref |  | 1 |  |  |  |  | 1 |  |  |
| Female | 0.46 | 0.28-0.64 | 1.58 | 1.32-1.90 | <0.001 | 0.43 | 0.26-0.60 | 1.53 | 1.29-1.82 | <0.001 |
| **Residence** |  |  |  |  |  |  |  |  |  |  |
| Inside ITS | Ref |  | 1 |  |  |  |  | 1 |  |  |
| Outside ITS | 0.17 | 0.00-0.34 | 1.19 | 1.00-1.41 | 0.045 | 0.16 | 0.00-0.32 | 1.17 | 1.00-1.38 | 0.045 |
| **Education** |  |  |  |  |  |  |  |  |  |  |
| Never attended school | Ref |  | 1 |  |  |  |  | 1 |  |  |
| Elementary | 0.24 | 0.02-0.45 | 1.27 | 1.02-1.57 | 0.030 | 0.22 | 0.02-0.42 | 1.25 | 1.02-1.52 | 0.030 |
| Preparatory and higher | 0.39 | 0.17-0.62 | 1.48 | 1.18-1.86 | 0.001 | 0.37 | 0.16-0.58 | 1.44 | 1.17-1.78 | 0.001 |
| **COVID-19 perceived severity** |  |  |  |  |  |  |  |  |  |  |
| COVID-19 is a serious infection that is spreading across the world |  |  |  |  |  |  |  |  |  |  |
| True | Ref |  | 1 |  |  |  |  | 1 |  |  |
| False | 0.51 | 0.16-0.86 | 1.66 | 1.17-2.37 | 0.005 | 0.47 | 0.15-0.80 | 1.61 | 1.16-2.23 | 0.005 |
| **COVID-19 Source of information** |  |  |  |  |  |  |  |  |  |  |
| Social Media |  |  |  |  |  |  |  |  |  |  |
| No | Ref |  | 1 |  |  |  |  | 1 |  |  |
| Yes | 0.25 | 0.08-0.41 | 1.28 | 1.09-1.51 | 0.003 | 0.23 | 0.08-0.38 | 1.26 | 1.08-1.46 | 0.003 |
| **Vaccination perceptions** |  |  |  |  |  |  |  |  |  |  |
| I think vaccines are safe |  |  |  |  |  |  |  |  |  |  |
| Agree | Ref |  | 1 |  |  |  |  |  |  |  |
| Neither agree nor disagree or Don't know | 0.56 | 0.35-0.78 | 1.76 | 1.42-2.17 | <0.001 | 0.53 | 0.33-0.72 | 1.69 | 1.39-2.06 | <0.001 |
| Disagree | 1.71 | 1.45-1.97 | 5.51 | 4.25-7.14 | <0.001 | 1.59 | 1.35-1.83 | 4.90 | 3.85-6.24 | <0.001 |
| **Intercept** | -3.71 | -4.59- - 2.83 | 0.02 | 0.01-0.06 | <0.001 | -3.51 | -3.59 - -3.43 | 0.03 | 0.03-0.03 | <0.001 |
| **Calibration and discrimination of models** |  |  |  |  |  |  |  |  |  |  |
|  |  | **95% CI** |  |  |  |  | **95% CI** |  |  |  |
| **C-statistic /Area under the curve** | 0.664 | 0.643-0.685 |  |  | 0.651 |  | 0.630-0.672 |  |  |  |
| **C-slope** | 1 | 0.869- 1.131 |  |  | 0.932 |  | 0.823-1.065 |  |  |  |
| **Calibration in the large** | 0 | -0.080-0.080 |  |  | 0.002 |  | -0.079-0.094 |  |  |  |

Similarly, in model 2, when perception around vaccine effectiveness was added as a predictor instead of vaccine safety, the C-statistic was 0.67 (95% CI: 0.65-0.69) indicating moderate discrimination and the model calibration curve also showed reasonably good agreement between predicted probability and observed incidence of COVID-19 vaccine refusal (Table S1). Variables that were predictors of COVID-19 vaccine refusal in model 2 were similar to those included in model 1 (Table S1).

Many of the coefficients of the predictors had the expected direction with the outcome: being female, residing outside an ITS, not perceiving COVID-19 as a severe illness, not perceiving vaccines as safe, and using of social media as a source of information for COVID-19.

### COVID-19 vaccine registration

Data on vaccine registration, collected in wave 4, showed that registration was low irrespective of intention to receive the vaccine as reported in wave 3, although the reasons differed between the two groups. Among participants in wave 4, 6.1% (n=141) registered to take the COVID-19 vaccine. Among those who intended to receive the vaccine 8.1% registered, among those who intended to refuse 2.7% registered, and among those undecided 4.8% registered. However, while the main reason for not registering among those who intended to receive the vaccine was not knowing how to register (42.6%); the main reason for not registering among those who intended to refuse in wave 3 was not wanting to receive the vaccine (46.5%). Although the proportion of those who were undecided in wave 3 was small, it is worth mentioning that their reasons for not registering on the vaccination platform were not knowing how to register (34.4%), not wanting to receive the vaccine (29%), or still not sure whether they want to take the vaccine (27.4%).

## Discussion

### Main findings

This study showed that although a large percentage of older Syrian refugees intended to receive the vaccine in wave 3 (61.3%), only a minute percentage had registered on the national platform to receive it in wave 4 (6%). Predictors of COVID-19 vaccine refusal were sex, age, education, living outside ITS, low perception of the severity of COVID-19, using social media as a source of information on COVID-19, and considering vaccines as not safe or effective. The primary reasons for COVID-19 vaccine refusal were: preference to follow preventive measures, concerns that the vaccine is too new, and belief that the vaccine is not essential. Vaccine registration on the MOPH Lebanon system was low with the two main reasons being unsure about how to register and not wanting to receive the vaccine.

### Findings in context

In contrast to other studies in the region, intention to accept the vaccine in January-April 2021 was higher in the Syrian refugee population than among the general adult population in Lebanon and the region. The study by Al Halabi and colleagues in 2021 showed that only 21.4% of Lebanese participants were willing to take the COVID-19 vaccine, 40.9% refused, and 37.7% were hesitant.^22^ In a multi-country study on vaccine acceptance, 44% of adults in Lebanon were willing to get vaccinated.^23^ Likewise, several Arab countries reported low vaccine acceptance rates including Kuwait (23.6%), Jordan (28.4%), and Saudi Arabia (31.8%).^24^.

Our findings have presented a number of reasons for vaccine refusal, which were similar to other studies. Participants in the oldest age groups were more likely to refuse COVID-19 vaccine than younger participants, an alarming finding given that older age is one of the main biological risk factors for COVID-19 severe illness and mortality. Similar to other studies in Lebanon^22^ but also in Qatar,^14^ France,^25^ UK,^26^ Australia,^27^ and the US,^28^ men reported higher intention to receive the vaccine as compared to women, with women in the UK reporting mistrust in the COVID-19 vaccine and higher concerns around its side effects.^26^ Hence, sex-sensitive public health interventions should be planned and implemented to address vaccine hesitancy among both male and female refugees.

Moreover, refugees living outside an ITS were more likely than those living inside an ITS to refuse receiving the COVID-19 vaccine. Humanitarian interventions usually target refugees living inside ITS since refugees inside these settlements are more identifiable. Our findings, however, highlight the need for more targeted and innovative interventions that are inclusive of and accessible to hard-to-reach populations including refugees in urban and residential areas.

In this study, using social media for information on COVID-19 was also a predictor of vaccine refusal among older Syrian refugees. Previous studies have shown that misinformation propagated through social media has shaped public opinion around COVID-19 and vaccination.^14^ Disseminating timely, evidence-based, culturally-sensitive and accessible information through diverse channels is highly needed for refugees. Community-based interventions engaging refugees, community leaders and religious figures can also play a significant role in building trust, disseminating accurate messages addressing COVID-19 and vaccine-related myths and misconceptions, and influencing positive social norms.^29 30^

A main finding revealed in our study is that, irrespective of initial vaccination intention, a large proportion of refugees in our sample did not register on the national platform for the COVID-19 vaccine. This goes in line with national figures showing that only 14.3% of Syrian refugees have registered for the COVID-19 vaccine up to January 2022.^17^ Therefore, to address the gap between intention to take the vaccine and actual uptake, practical aspects such as having access to the online platform or knowing how to register need to be addressed. In this regard, more assistance should be provided to refugees in community-based interventions to help them register online for the COVID-19 vaccine. Among those who did not intend to take the vaccine, the most reported reason for not registering was not wanting to receive the vaccine, while the main reason for not registering among those who intended to take it was related to barriers in the registration process.

Vaccine uptake has been shown to be influenced by several contextual, individual, and vaccine factors related to Complacency, Convenience and Confidence (3C model).^31^ In this study, perceptions around COVID-19 severity, and about vaccine safety and effectiveness were predictors of COVID-19 vaccine refusal among older Syrian refugees. Future vaccination interventions and communication strategies should be informed by (1) addressing complacency barriers related to perceived susceptibility and severity of COVID-19, (2) convenience barriers including vaccine accessibility, availability and affordability and (3) confidence barriers related to vaccine safety and effectiveness and general mistrust in the system. The 3C model is relevant to the refugee populations as previous studies have identified barriers to vaccination as including lacking legal documentation, fear of deportation through data sharing with immigration authorities or security checks, mistrust in authorities, and concerns regarding vaccine affordability, accessibility, safety and effectiveness.^32^ The COVID-19 pandemic has further increased social exclusion and aggravated issues of mistrust in the public health system impacting willingness to vaccinate. To tackle these concerns, information on the process of COVID-19 vaccine registration and around personal data sharing need to be clearly and effectively communicated. In addition, innovative public health interventions, such as mobile vaccine clinics, to overcome barriers related to security checks, mistrust in authorities, and concerns regarding vaccine affordability, and accessibility.

### Strengths and limitations

This study is the largest longitudinal study on Syrian refugees in Lebanon during the COVID-19 pandemic and one of the first to present evidence on COVID-19 vaccine acceptance and examine predictors of COVID-19 vaccine refusal among refugees in the region. Nonetheless, it is representative of beneficiaries of a humanitarian organization and not of all Syrian refugees in Lebanon. The study has several limitations: the study outcome, intention to vaccinate, was asked during the early phases of the vaccine campaign in Lebanon and thus intention data may be subjective and not reflect actual vaccination uptake had the data been collected months later as the vaccine became more readily available. Since data collection of wave 4, there have been many public health initiatives by international NGOs to increase vaccine acceptance, registration on the MOPH platform, and actual uptake of the vaccine among refugees in Lebanon.

## Conclusion

The inclusion of refugees in vaccination programs regardless of their legal and documentation status is vital to ensure equitable vaccination, and the prevention of COVID-19 related illness and death. Findings from this study highlight the need to enhance vaccine acceptance and uptake among older Syrian refugees, and address barriers to vaccine registration. A better understanding of predictors of COVID-19 vaccine refusal promises to inform better humanitarian programming and health promotion targeting now and in the face of future health crises. Building culturally- and gender-sensitive immunization programs that meet the specific needs of refugees and inclusive communication sources and messages to re-establish trust and counter existing concerns, myths and misinformation particularly in relation to vaccine safety and effectiveness are required.

## Data Availability

All data produced in the present study are available upon reasonable request to the authors

## Contributors

SA, HG, and SM conceptualized the study and the survey design. SA, SM, MH and NS contributed to data collection and analysis. NS contributed to the literature search and wrote the first draft of the paper; following drafts were reviewed and revised by SA and SM. The underlying survey data were verified by SM and NS. SM supervised NS and BZ throughout the project. All authors have reviewed and approved the final version of the paper.

