## Supplementary material for "Predicting intention to vaccinate against COVID-19 in older Syrian refugees in Lebanon: findings from a multi-wave study": Table S1

**Supplementary Materials**

***Table S1. Multivariable model for predicting COVID-19 vaccine refusal using vaccine effectiveness***

|  | **Apparent Model** | | | | | **Model adjusted by bootstrap shrinkage** | | | | |
| --- | --- | --- | --- | --- | --- | --- | --- | --- | --- | --- |
|  | **Parameter estimate** | **95%CI** | **Odds Ratio** | **95% CI** | **P-value** | **Parameter estimate** | **95%CI** | **Odds Ratio** | **95% CI** | **P-value** |
| ***Age*** 0.02 | | 0.01-0.03 | 1.02 | 1.01-1.03 | 0.003 | 0.01 | 0.00-0.02 | 1.01 | 1.01-1.03 | 0.003 |
| ***Sex*** | |  |  |  |  |  |  |  |  |  |
| *Male* | Ref |  | 1 |  |  |  |  | 1 |  |  |
| *Female* | 0.47 | 0.28-0.65 | 1.60 | 1.33-1.92 | <0.001 | 0.44 | 0.26-0.61 | 1.55 | 1.30-1.83 | <0.001 |
| ***Residence*** |  |  |  |  |  |  |  |  |  |  |
| *Inside ITS* | Ref |  | 1 |  |  |  |  | 1 |  |  |
| *Outside ITS* | 0.17 | -0.00-0.34 | 1.18 | 1.00-1.40 | 0.050 | 0.16 | -0.00-0.32 | 1.17 | 1.00-1.37 | 0.050 |
| ***Education*** |  |  |  |  |  |  |  |  |  |  |
| *Never attended school* | Ref |  | 1 |  |  |  |  | 1 |  |  |
| *Elementary* | 0.22 | 0.01-0.43 | 1.25 | 1.01-1.54 | 0.044 | 0.20 | 0.01-0.40 | 1.23 | 1.01-1.50 | 0.044 |
| *Preparatory and higher* | 0.40 | 0.17-0.62 | 1.49 | 1.18-1.87 | 0.001 | 0.37 | 0.16-0.58 | 1.45 | 1.17-1.79 | 0.001 |
| ***COVID-19 perceived severity*** |  |  |  |  |  |  |  |  |  |  |
| ***COVID-19 is a serious infection that is spreading across the world*** |  |  |  |  |  |  |  |  |  |  |
| *True* | Ref |  | 1 |  |  |  |  | 1 |  |  |
| *False* | 0.51 | 0.16-0.86 | 1.67 | 1.17-2.37 | 0.005 | 0.48 | 0.15-0.80 | 1.61 | 1.16-2.23 | 0.005 |
| ***COVID-19 Source of information*** |  |  |  |  |  |  |  |  |  |  |
| ***Social Media*** |  |  |  |  |  |  |  |  |  |  |
| *No* | Ref |  | 1 |  |  |  |  | 1 |  |  |
| *Yes* | 0.26 | 0.10-0.43 | 1.30 | 1.11-1.54 | 0.002 | 0.25 | 0.09-0.40 | 1.28 | 1.10-1.49 | 0.002 |
| ***Vaccination perceptions*** |  |  |  |  |  |  |  |  |  |  |
| ***I think vaccines are effective*** |  |  |  |  |  |  |  |  |  |  |
| *Agree* | Ref |  | 1 |  |  |  |  |  |  |  |
| *Neither agree nor disagree or Don’t know* | 0.63 | 0.43-0.84 | 1.88 | 1.53-2.32 | <0.001 | 0.59 | 0.40-0.78 | 1.80 | 1.49-2.19 | <0.001 |
| *Disagree* | 1.89 | 1.61-2.18 | 6.65 | 5.00-8.85 | <0.001 | 1.76 | 1.50-2.03 | 5.83 | 4.47-7.61 | <0.001 |
| ***Intercept*** | -3.74 | -4.62- -2.86 | 0.02 | 0.01-0.06 | <0.001 | -3.53 | -3.61- -3.45 | 0.03 | 0.03-0.03 | <0.001 |
| ***Calibration and discrimination of models*** |  |  |  |  |  |  |  |  |  |  |
|  |  | **95% CI** |  |  |  |  | **95% CI** |  |  |  |
| ***C-statistic /Area under the curve*** | 0.669 | (0.648-0.690) |  |  | 0.656 |  | 0.636-0.677 |  |  |  |
| ***C-slope*** | 1 | (0.872-1.128) |  |  | 0.931 |  | 0.817-1.055 |  |  |  |
| **Calibration in the large** | 0 | (-0.080-0.080) |  |  | 0.002 |  | -0.076-0.091 |  |  |  |
